# Human Safety, Tolerability, and Pharmacokinetics of a Novel Broad-Spectrum Oral Antiviral Compound, Molnupiravir, with Activity Against SARS-CoV-2

**DOI:** 10.1101/2020.12.10.20235747

**Authors:** Wendy P. Painter, Wayne Holman, Jim A. Bush, Firas Almazedi, Hamzah Malik, Nicola C. J. E. Eraut, Merribeth J. Morin, Laura J. Szewczyk, George R. Painter

## Abstract

Molnupiravir, EIDD-2801/MK-4482, the prodrug of the ribonucleoside analog ß-d-N4-hydroxycytidine (NHC), has activity against a number of RNA viruses including severe acute respiratory syndrome coronavirus 2, severe acute respiratory syndrome coronavirus, Middle East respiratory syndrome coronavirus, seasonal and pandemic influenza viruses, and respiratory syncytial virus.

Single and multiple doses of molnupiravir were evaluated in this first-in-human, phase 1, randomized, double-blind, placebo-controlled study in healthy volunteers, which included evaluation of the effect of food on pharmacokinetics.

EIDD-1931 appeared rapidly in plasma, with a median time of maximum observed concentration of 1.00 to 1.75 hours, and declined with a geometric half-life of approximately 1 hour, with a slower elimination phase apparent following multiple doses or higher single doses (7.1 hours at the highest dose tested). Mean maximum observed concentration and area under the concentration versus time curve increased in a dose-proportional manner, and there was no accumulation following multiple doses. When administered in a fed state, there was a decrease in the rate of absorption, but no decrease in overall exposure.

Molnupiravir was well tolerated. Fewer than half of subjects reported an adverse event, the incidence of adverse events was higher following administration of placebo, and 93.3% of adverse events were mild. One discontinued early due to rash. There were no serious adverse events and there were no clinically significant findings in clinical laboratory, vital signs, or electrocardiography. Plasma exposures exceeded expected efficacious doses based on scaling from animal models; therefore, dose escalations were discontinued before a maximum tolerated dose was reached.

**Clinical trial identifier:** This study was registered at ClinicalTrials.gov with the identifier NCT04392219.

## Introduction

A novel coronavirus, originally identified in Wuhan City, China, was reported to the World Health Organization on 31 December 2019 (1) and the associated disease has subsequently become a worldwide pandemic. An effective antiviral therapeutic has since been intensively sought.

The causative agent of the current pandemic, severe acute respiratory syndrome coronavirus 2 (SARS-CoV-2), is a sister clade to severe acute respiratory syndrome coronavirus (SARS-CoV), the causative agent of severe acute respiratory syndrome (SARS). SARS emerged in Guangdong, China in 2002, causing over 700 deaths among over 8000 cases. The severity of this epidemic in 2002-2003 was limited by the relatively low transmissibility of SARS-CoV and association of peak viremia with clinical symptoms rather than a prodromal phase (2); however, the epidemic demonstrated the potential for the rapid spread of an infectious disease in a highly connected and interdependent world. This risk was emphasized when another novel coronavirus, the Middle East respiratory syndrome coronavirus (MERS-CoV), the causative agent of Middle East respiratory syndrome (MERS), emerged in 2012 and resulted in a localized epidemic in 2014.

SARS-CoV-2 has now established itself globally in the human population. Thus, there is an urgent need for an effective, oral antiviral that can be manufactured and broadly distributed on a scale adequate to meet global need. Such a therapy could prevent deaths, reduce disease severity, reduce hospital admission times and thus lessen stress on healthcare resources, potentially limit the spread of SARS-CoV-2, and ameliorate the global and economic disruption that was initiated by the Coronavirus disease-2019 (COVID-19) pandemic.

COVID-19 may be asymptomatic, mild, severe, life-threatening, or fatal. While the majority of cases can be managed in an outpatient setting, patients with risk factors including advanced age, cardiovascular and pulmonary comorbidities, diabetes, obesity, chronic kidney disease, and cancer are at higher risk of developing more severe disease that may require hospitalization or may result in death and severe morbidity. Asymptomatic carriers and those with mild disease may spread the disease to those at high risk and may subsequently suffer less common and emerging medium-to-longer term sequelae, such as multi-system inflammatory syndrome in children or prolonged cardiopulmonary compromise in otherwise healthy adults (3). Healthcare utilization stresses in regions with outbreaks, quarantines, school closures, and isolation of the elderly may create further disruption and morbidity because of effects on the normal running of hospital services, including cancer referrals and management of chronic conditions. The overall fatality rate is likely higher than for influenza, but estimates are still changing based on emerging data (4). As with other viral respiratory diseases, fatalities are heavily skewed toward older patients and those with risk factors; children are rarely symptomatic and rarely have serious illness.

COVID-19 is primarily a respiratory disease, but many extrapulmonary effects and complications have been described. Cough and fever are the most frequent symptoms, followed by myalgia, headache, dyspnea, sore throat, diarrhea, nausea/vomiting, and anosmia or ageusia (5). These initial COVID-19 symptoms are not easily distinguished from other respiratory infections such as influenza; therefore, a broad-spectrum treatment would be advantageous for empiric therapy. Complications of COVID-19 include respiratory failure, cardiovascular events, thromboembolic events, and inflammatory damage (6), and respiratory sequelae are anticipated in patients with severe disease (7). Thus, administration of a potent, direct-acting antiviral at various stages of viral replication may reduce not only acute disease symptoms and transmission, but also complications and sequelae of untreated progressive disease.

Molnupiravir (also known as EIDD-2801/MK-4482) is a prodrug of the ribonucleoside analog ß -d-N4-hydroxycytidine (EIDD-1931 [NHC]), which is phosphorylated intracellularly to the active 5′-triphosphate. Molnupiravir has demonstrated the potential to treat infections caused by several RNA viruses, including pandemic-capable coronaviruses and influenza viruses, and encephalitic alpha viruses such as Venezuelan, Eastern, and Western equine encephalitis viruses in nonclinical models. The primary mechanism of action of molnupiravir against RNA viruses is viral error catastrophe (8, 9), a concept that is predicated on increasing the viral mutation rate beyond a biologically tolerable threshold, resulting in impairment of viral fitness and leading to viral extinction. Molnupiravir has demonstrated *in vitro* activity against SARS-CoV-2 in human airway epithelial cell cultures. Prophylactic and therapeutic administration of molnupiravir to mice infected with SARS-CoV or MERS-CoV improved pulmonary function, and reduced virus titer and body weight loss (10). In the ferret influenza model, treatment of pandemic influenza A virus with molnupiravir resulted in reduced viral shedding and inflammatory cellular infiltrates in nasal lavages, with a normal humoral antiviral response (11).

Here we report the results of a first-in-human, phase 1, randomized, double-blind, placebo-controlled study to determine the safety, tolerability, and pharmacokinetics of single and multiple ascending oral doses of molnupiravir in healthy subjects. A randomized, open-label, crossover evaluation in the fed (high fat) and fasted states was also conducted to assess the effect of food on the pharmacokinetics of single doses of molnupiravir.

## Results

Eligible subjects were randomized in a 3:1 ratio to either study drug or placebo in the single- and multiple-ascending-dose parts of the study. Each cohort comprised 8 subjects, with single oral doses of 50 to 1600 mg administered in the single-ascending-dose part and twice-daily (BID) doses of 50 to 800 mg administered for 5.5 days in the multiple-ascending-dose part. Subjects were followed for 14 days following completion of dosing for assessments of safety, tolerability, and pharmacokinetics. Subjects in the food-effect evaluation were randomized in a 1:1 ratio to either 200 mg molnupiravir in the fed state followed by 200 mg molnupiravir in the fasted state, or vice versa, with a 14-day washout period between doses. A capsule formulation was used in all parts of the study, with the exception of single ascending doses ≤800 mg, where an oral solution formulation was used.

### Disposition

Sixty-four subjects received a single dose of between 50 and 1600 mg molnupiravir or placebo; 55 subjects received between 50 and 800 mg molnupiravir or placebo BID for 5.5 days; and 10 subjects received a single dose of 200 mg molnupiravir in the fed state followed by a single dose of 200 mg molnupiravir in the fasted state after a washout period of 14 days, or vice versa. Additionally, 1 subject in the multiple-ascending-dose part received 800 mg molnupiravir BID for 3 days, but was discontinued from dosing by the investigator on Day 4. All subjects completed the protocol-specified study procedures and assessments.

### Demography

Subjects were aged between 19 and 60 years, with a mean body mass index between 24.4 and 25.4 kg/m^2^ (Table 1). The majority of subjects were white and male. There were no other notable differences in subject demography between cohorts, except for age, where the mean age was higher in the food-effect evaluation cohort, the 50-mg molnupiravir single-dose cohort, and in the 100-mg molnupiravir multiple-dose cohort (data not shown).

**Table 1.**
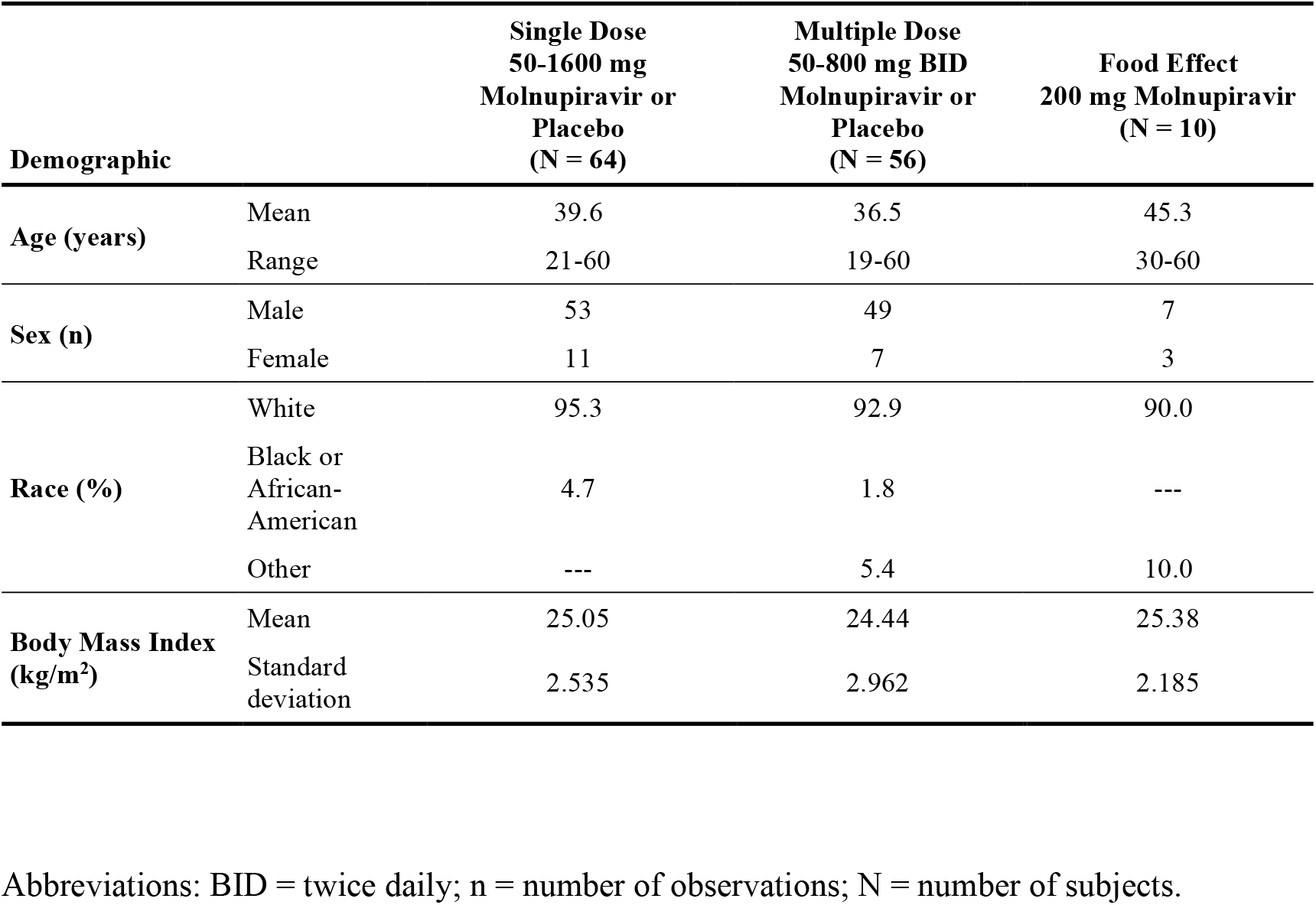
Demography

### Tolerability

Adverse events were graded using the Division of Microbiology and Infectious Diseases (DMID) toxicity grading, dated March 2014.**(i) Single ascending doses.** Overall, 37.5% of subjects reported an adverse event (Table 2). There were no apparent dose-related trends, with a greater proportion of subjects reporting adverse events following administration of placebo (43.8%) than following administration of molnupiravir (35.4%). Only 1 moderate adverse event (headache; Grade 2) was reported following administration of molnupiravir, which occurred at the 400-mg dose level. One subject also reported moderate adverse events (nausea and headache; Grade 2) following administration of placebo. No severe (Grade 3) adverse events were reported. The most frequently reported adverse event was headache, which was reported by 18.8% of subjects who were administered placebo and 12.5% of subjects who were administered molnupiravir. **(ii) Multiple ascending doses.** Overall, 44.6% of subjects reported an adverse event (Table 3). There were no apparent dose-related trends, with a greater proportion of subjects reporting adverse events following administration of placebo (50.0%) than following administration of molnupiravir (42.9%). With the exception of 1 subject who reported moderate (Grade 2) events of oropharyngeal pain, pain in extremity, and influenza-like illness, all adverse events were mild (Grade 1) in severity. The most frequently reported adverse event was diarrhea, which was reported by 7.1% of subjects who were administered molnupiravir and 7.1% of subjects who were administered placebo. One subject discontinued study drug administration on Day 4 because of an adverse event of mild, truncal, maculopapular, pruritic rash following multiple BID doses of 800 mg molnupiravir, which was considered by the investigator to be related to the study drug. Following discontinuation, the subject was administered potent topical steroid treatment and anti-histamines, and pruritis and rash had both resolved within 18 days.**(iii) Food-effect evaluation.**Three subjects in the food-effect evaluation each reported 1 adverse event, all of which were mild (Grade 1) in severity.

There were no serious adverse events reported in this study and there were no trends of increased frequency or severity of adverse events with higher doses of molnupiravir.

**Table 2.** Treatment-emergent adverse events (50-1600 mg molnupiravir single ascending doses)

|  | Placebo<br>(N = 16) | 50 mg<br>(N = 6) | 100 mg<br>(N = 6) | 200 mg<br>(N = 6) | 400 mg<br>(N = 6) | 600 mg<br>(N = 6) | 800 mg<br>(N = 6) | 1200 mg<br>(N = 6) | 1600 mg<br>(N = 6) |
| --- | --- | --- | --- | --- | --- | --- | --- | --- | --- |
| **Overall (n, [nE])** | 7 [12] | 2 [4] | --- | 3 [4] | 3 [5] | 4 [7] | 2 [5] | 1 [2] | 2 [8] |
| **Severity** |  |  |  |  |  |  |  |  |  |
| Mild<br>(Grade 1) | 7 [10] | 2 [4] | --- | 3 [4] | 3 [4] | 4 [7] | 2 [5] | 1 [2] | 2 [8] |
| Moderate<br>(Grade 2) | 1 [2] | --- | --- | --- | 1 [1] | --- | --- | --- | --- |
| Severe<br>(Grade 3) | --- | --- | --- | --- | --- | --- | --- | --- | --- |
| **Related** | 2 [4] | 1 [1] | --- | 2 [3] | --- | 1 [1] | --- | --- | --- |
| **Preferred Terms Reported By More Than 1 Subject (n)** |  |  |  |  |  |  |  |  |  |
| Headache | 3 | 1 | --- | --- | 1 | 3 | 1 | --- | --- |
| Catheter site pain* | 1 | --- | --- | --- | 1 | 1 | --- | --- | --- |
| Nausea | 1 | --- | --- | 1 | --- | 1 | --- | --- | --- |
| Rhinorrhoea | 1 | --- | --- | --- | --- | --- | 1 | 1 | --- |
| Back pain | --- | --- | --- | --- | --- | 1 | 1 | --- | --- |
| Feeling hot | --- | --- | --- | --- | 2 | --- | --- | --- | --- |
| Pain in extremity | 1 | 1 | --- | --- | --- | --- | --- | --- | --- |

Abbreviations: n = number of subjects with an adverse event; nE = number of adverse events; N = number of subjects.
\* venous cannula site pain

**Table 3.** Treatment-emergent adverse events (50-800 mg molnupiravir twice-daily multiple ascending doses)

|  | **Placebo**<br>**BID**<br>**(N = 14)** | **50 mg**<br>**BID**<br>**(N = 6)** | **100 mg**<br>**BID**<br>**(N = 6)** | **200 mg**<br>**BID**<br>**(N = 6)** | **300 mg**<br>**BID**<br>**(N = 6)** | **400 mg**<br>**BID**<br>**(N = 6)** | **600 mg**<br>**BID**<br>**(N = 6)** | **800 mg**<br>**BID**<br>**(N = 6)** |
| --- | --- | --- | --- | --- | --- | --- | --- | --- |
| **Overall (n, [nE])** | 7 [11] | 2 [2] | 3 [3] | 3 [9] | 2 [3] | 3 [5] | 2 [2] | 3 [5] |
| **Severity** |  |  |  |  |  |  |  |  |
| Mild<br>(Grade 1) | 7 [11] | 2 [2] | 3 [3] | 3 [6] | 2 [3] | 3 [5] | 2 [2] | 3 [5] |
| Moderate<br>(Grade 2) | --- | --- | --- | 1 [3] | --- | --- | --- | --- |
| Severe<br>(Grade 3) | --- | --- | --- | --- | --- | --- | --- | --- |
| **Related** | 3 [4] | --- | --- | 2 [3] | 1 [1] | --- | 1 [1] | 3 [4] |
| **Preferred Terms Reported By More Than 1 Subject (n)** |  |  |  |  |  |  |  |  |
| Diarrhoea | 1 | --- | --- | 1 | --- | --- | 1 | 1 |
| Back pain | --- | --- | 2 | --- | 1 | --- | --- | --- |
| Headache | --- | --- | 1 | --- | 2 | --- | --- | --- |
| Somnolence | 2 | --- | --- | --- | --- | 1 | --- | --- |

Abbreviations: BID = twice daily; n = number of subjects with an adverse event; nE = number of adverse events; N = number of subjects.

### Safety

There were no clinically significant findings or dose-related trends in clinical laboratory, vital signs, and electrocardiogram data. The dose-limiting toxicity in one Investigational New Drug-enabling study (in the dog, the most sensitive species) was bone marrow toxicity and included reversible reductions in platelet counts; however, no clinically significant changes in hematological parameters were seen in this study.

Dose escalations were discontinued before a maximum tolerated dose was reached because plasma exposures that were expected to be efficacious based on scaling from animal models of seasonal and pandemic influenza were exceeded (11).

### Pharmacokinetics

#### (i) Single ascending doses

Concentrations of molnupiravir were generally not quantifiable at doses up to 800 mg, with the exception of the 0.25-hour timepoint after doses of 600 and 800 mg, where concentrations were quantifiable in 5 and 4 subjects, respectively, and the 0.5-hour timepoint after a dose of 800 mg, where concentrations were quantifiable in all subjects. At doses of 1200 and 1600 mg, concentrations of molnupiravir were quantifiable at 1 or more timepoints between 0.25 and 1.5 hours postdose in all subjects. Molnupiravir pharmacokinetic parameters were not calculable for doses ≤400 mg; however, at doses ≥600 mg, maximum observed concentration (C_max_), time of C_max_ (t_max_), and time of last quantifiable concentration were calculable. Following administration of between 600 and 1600 mg molnupiravir, values of mean C_max_ were up to 13.2 ng/mL and values of median t_max_ were between 0.25 and 0.75 hours (data not shown). It should be noted that molnupiravir concentrations represented only approximately 0.2% of EIDD-1931 concentrations and t_max_ of molnupiravir occurred at the first sampling timepoint for the 600-mg dose level, and therefore C_max_ may have been underestimated. At doses of ≥800 mg, trace amounts of molnupiravir were detected in the urine, which represented approximately 0.002% of the dose (data not shown).

Following oral administration of molnupiravir at doses up to 800 mg, EIDD-1931 appeared rapidly in plasma, with a median t_max_ of 1.00 hour postdose in all dose cohorts, after which plasma concentrations declined in an essentially monophasic manner with geometric mean terminal elimination half-lives (t_1/2_) of between 0.910 and 1.29 hours postdose (Table 4 and Figure 1). However, at doses of 1200 and 1600 mg, median t_max_ was delayed, with median t_max_ occurring at 1.75 and 1.50 hours, respectively. Plasma concentrations at doses of 1200 and 1600 mg were quantifiable, along with a second slower elimination phase, where mean t_1/2_ was longer with values of 1.81 and 4.59 hours, respectively.

**Table 4.** Pharmacokinetic parameters of EIDD-1931 (50-1600 mg molnupiravir single ascending doses) Geometric means (percentage coefficient of variation) are presented, with the exception of t_max_ where medians (minimum – maximum) are presented.

| Parameter | 50 mg<br>(N = 6) | 100 mg<br>(N = 6) | 200 mg<br>(N = 6) | 400 mg<br>(N = 6) | 600 mg<br>(N = 6) | 800 mg<br>(N = 6) | 1200 mg<br>(N = 6) | 1600 mg<br>(N = 6) |
| --- | --- | --- | --- | --- | --- | --- | --- | --- |
| **AUC<sub>last</sub> (ng.h/mL)** | 415 (27.4) | 917<br>(27.5) | 1810<br>(20.0) | 4000<br>(20.2) | 6120<br>(21.6) | 8720<br>(10.4) | 13800<br>(11.7) | 20700<br>(31.4) |
| **AUC<sub>inf</sub> (ng.h/mL)** | 432 (26.5) | 932<br>(27.0) | 1830<br>(19.6) | 4010<br>(20.2) | 6130<br>(21.4) | 8740<br>(10.4) | 13800<br>(11.8) | 20700<br>(31.4) |
| **C<sub>max</sub> (ng/mL)** | 223 (46.2) | 454<br>(42.2) | 926<br>(12.6) | 1850<br>(22.7) | 2720<br>(27.0) | 3640<br>(13.4) | 4500<br>(17.9) | 6350<br>(20.6) |
| **t<sub>max</sub> (h)** | 1.00<br>(0.517 –<br>1.00) | 1.00<br>(0.500 –<br>1.50) | 1.00<br>(0.500 –<br>1.00) | 1.00<br>(0.500 –<br>1.00) | 1.00<br>(1.00 –<br>1.00) | 1.00<br>(0.500 –<br>1.00) | 1.75<br>(1.00 –<br>2.50) | 1.50<br>(1.00 –<br>2.00) |
| **t<sub>1/2</sub> (h)** | 0.945<br>(12.1) | 0.907<br>(10.1) | 1.02<br>(16.4) | 1.03<br>(8.86) | 1.06<br>(10.3) | 1.29<br>(7.10) | 1.81<br>(73.5) | 4.59<br>(71.6) |
| **Ae<sub>0-24</sub> (mg)** | 0.323<br>(53.6) | 1.03<br>(68.1) | 1.51<br>(86.2) | 5.47<br>(108) | 14.4<br>(47.7) | 18.0<br>(14.7) | 53.7<br>(41.4) | 84.4<br>(29.9) |
| **Fe<sub>0-24</sub> (%)** | 0.820<br>(53.6) | 1.31<br>(68.1) | 0.958<br>(86.2) | 1.74<br>(108) | 3.05<br>(47.7) | 2.86<br>(14.7) | 5.69<br>(41.4) | 6.70<br>(29.9) |

Abbreviations: Ae<sub>0-24</sub> = amount of the dose administered recovered in urine from time 0 to 24 hours postdose; AUC<sub>inf</sub> = area under the plasma concentration-time curve from time 0 extrapolated to infinity; AUC<sub>last</sub> = area under the plasma concentration-time curve from time 0 to the last measurable non-zero concentration; C<sub>max</sub> = maximum observed concentration; Fe<sub>0-24</sub> = percentage of the dose administered recovered in urine from time 0 to 24 hours postdose; N = number of subjects; t<sub>1/2</sub> = apparent terminal elimination half-life; t<sub>max</sub> = time of the maximum observed concentration.

**Figure 1.**
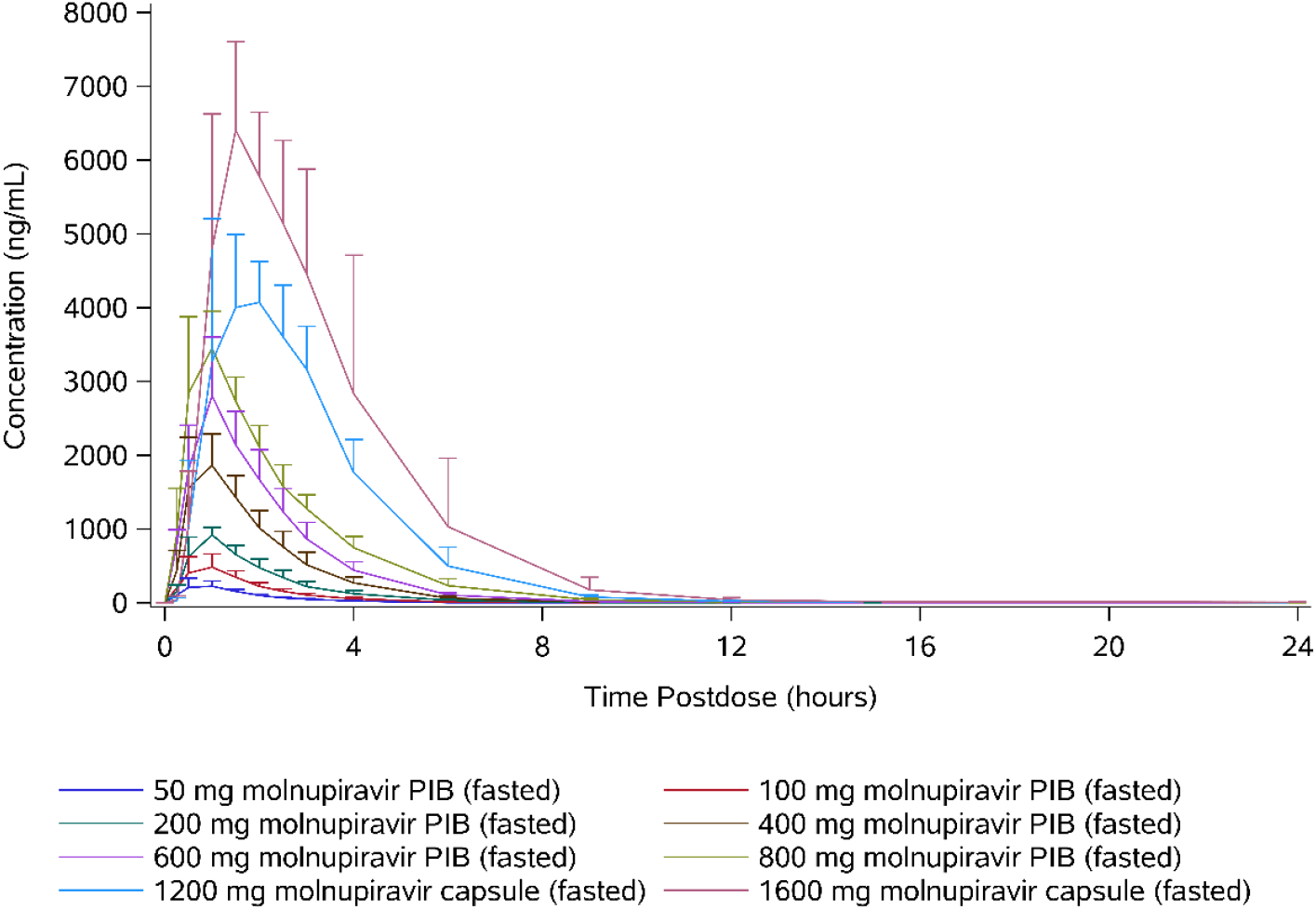
Arithmetic mean plasma concentrations of EIDD-1931 (50-1600 mg molnupiravir single ascending doses).

The plasma concentration-time profiles were generally well defined, with the percentage of area under the plasma concentration-time curve from time 0 extrapolated to infinity (AUC_inf_) that was extrapolated being <10% for all subjects. When assessed using a power model (ln[parameter] = intercept + slope x ln[dose] + random error), mean C_max_ increased in a dose-proportional manner, with the 90% confidence interval containing unity. Similarly, mean AUC_inf_ increased in an approximately dose-proportional manner; however, the lower bound of the 90% confidence interval was slightly above unity (Table 5).

**Table 5.** Dose proportionality of EIDD-1931 (50-800 mg molnupiravir single ascending doses)Abbreviations: AUC_inf_ = area under the plasma concentration-time curve from time 0 extrapolated to infinity; AUC_last_ = area under the plasma concentration-time curve from time 0 to the last measurable non-zero concentration; C_max_ = maximum observed concentration; n = number of observations.

| Parameter | n | Slope (90%<br>Confidence<br>Interval) | Between-subject<br>Geometric<br>Coefficient of<br>Variation | Lack of Fit<br>P-value |
| --- | --- | --- | --- | --- |
| AUC<sub>inf</sub> (ng.h/mL) | 36 | 1.07 (1.02, 1.13) | 21.5 | 0.9724 |
| AUC<sub>last</sub> (ng.h/mL) | 36 | 1.09 (1.03, 1.15) | 21.9 | 0.9750 |
| C<sub>max</sub> (ng/mL) | 36 | 1.01 (0.927, 1.08) | 29.8 | 0.9996 |

The amount of EIDD-1931 excreted in urine from time 0 to 24 hours postdose (Ae_0-24_) increased supraproportionally with dose, and there was a similar trend for apparent clearance (CL_R_) to increase. Between 0.820% (at the 50-mg dose level) and 6.70% (at the 1600-mg dose level) of the dose was excreted in urine as EIDD-1931, and the majority of the total amount was generally excreted within the first 4 hours postdose.

#### (ii) Multiple ascending doses

Concentrations of molnupiravir were generally not quantifiable at doses ≤400 mg BID and pharmacokinetic parameters were not calculable. Concentrations of molnupiravir were quantifiable in 4 subjects at either 0.5 or 1 hour postdose on Day 1 and in 3 subjects at 0.5 hours postdose on Day 6 at the 600-mg BID dose level. At the 800-mg dose level, concentrations of molnupiravir were quantifiable from all except 1 subject at 0.5 hours postdose on Days 1 and 6, but at no other timepoints, consistent with single ascending doses.

Following oral administration of molnupiravir, EIDD-1931 appeared rapidly in plasma, with a median t_max_ in all dose cohorts of between 1.00 and 1.75 hours postdose across both Days 1 and 6 (Table 6 and Figure 2). For all dose levels, plasma concentrations declined in an essentially monophasic manner on Day 1, with mean t_1/2_ ranging from 0.918 to 1.18 hours. Similarly, plasma concentrations declined in an essentially monophasic manner on Day 6 for subjects at dose levels ≤200 mg BID and for the majority of subjects at the 300- and 400-mg BID dose levels. Contrastingly, for 1 subject at each of the 300- and 400-mg dose levels and for all subjects at the 600- and 800-mg BID dose levels, there was the emergence of a second, slower elimination phase on Day 6, which was reflected in an increase in the mean t_1/2_ with increasing dose at doses ≥200 mg. Of note, at the 600-mg BID dose level, the lack of a clearly defined terminal elimination phase confounded the evaluation of t_1/2_ for the majority of subjects. At the 800-mg BID dose level, the mean t_1/2_ was 7.08 hours.

**Table 6.** Pharmacokinetic parameters of EIDD-1931 (50-800 mg molnupiravir twice-daily multiple ascending doses) Geometric means (percentage coefficient of variation) are presented, with the exception of t_max_ where medians (minimum – maximum) are presented.

| Parameter | Day 1 |  |  |  |  |  |  | Day 6 |  |  |  |  |  |  |
| --- | --- | --- | --- | --- | --- | --- | --- | --- | --- | --- | --- | --- | --- | --- |
|  | 50 mg<br>(N = 6) | 100 mg<br>(N = 6) | 200 mg<br>(N = 6) | 300 mg<br>(N = 6) | 400 mg<br>(N = 6) | 600 mg<br>(N = 6) | 800 mg<br>(N = 6) | 50 mg<br>(N = 6) | 100 mg<br>(N = 6) | 200 mg<br>(N = 6) | 300 mg<br>(N = 6) | 400 mg<br>(N = 6) | 600 mg<br>(N = 6) | 800 mg<br>(N = 5) |
| **AUC<sub>τ</sub>**<br>**(ng.h/mL)** | 461<br>(15.7) | 854<br>(19.8) | 1660<br>(15.3) | 3080<br>(17.3) | 3800<br>(19.5) | 6110<br>(26.9) | 8190<br>(21.5) | 432<br>(14.9) | 968<br>(15.3) | 1730<br>(25.2) | 2960<br>(16.2) | 3710<br>(21.6) | 7110<br>(28.2) | 8330<br>(17.9) |
| **AUC<sub>last</sub>**<br>**(ng.h/mL)** | 444<br>(17.3) | 835<br>(19.9) | 1640<br>(15.5) | 3080<br>(17.4) | 3790<br>(19.5) | 6110<br>(26.9) | 8180<br>(21.5) | 414<br>(16.2) | 947<br>(15.7) | 1720<br>(26.0) | 2980<br>(16.3) | 3730<br>(21.6) | 7250<br>(28.1) | 8450<br>(18.5) |
| **AUC<sub>inf</sub>**<br>**(ng.h/mL)** | 461<br>(15.7) | 855<br>(19.8) | 1660<br>(15.3) | 3090<br>(17.4) | 3800<br>(19.5) | 6680*<br>(17.6) | 8200<br>(21.6) | --- | --- | --- | --- | --- | --- | --- |
| **C<sub>max</sub>**<br>**(ng/mL)** | 223<br>(19.4) | 395<br>(18.5) | 766<br>(16.3) | 1280<br>(15.2) | 1530<br>(23.2) | 2160<br>(31.4) | 2770<br>(13.3) | 188<br>(8.67) | 434<br>(14.0) | 742<br>(32.1) | 1100<br>(20.6) | 1470<br>(20.9) | 2240<br>(20.9) | 2970<br>(16.8) |
| **t<sub>max</sub> (h)** | 1.00<br>(1.00 –<br>1.00) | 1.25<br>(1.00 –<br>2.03) | 1.50<br>(1.00 –<br>1.50) | 1.50<br>(1.00 –<br>1.50) | 1.50<br>(1.00 –<br>2.00) | 1.75<br>(1.00 –<br>6.00) | 1.75<br>(1.50 –<br>2.50) | 1.00<br>(1.00 –<br>1.50) | 1.25<br>(1.00 –<br>1.50) | 1.50<br>(0.500 –<br>1.50) | 1.50<br>(1.00 –<br>2.00) | 1.50<br>(1.00 –<br>1.50) | 1.75<br>(1.50 –<br>2.50) | 1.50<br>(1.00 –<br>2.02) |
| **MR<sub>Cmax</sub>** | NC | NC | NC | NC | NC | 329*<br>(87.7) | 512<br>(30.6) | NC | NC | NC | NC | NC | NC | 413†<br>(28.1) |
| **t<sub>1/2</sub> (h)** | 0.937<br>(14.0) | 0.918<br>(9.08) | 0.960<br>(10.4) | 1.09<br>(17.7) | 1.05<br>(13.1) | 1.16*<br>(3.50) | 1.18<br>(7.28) | 0.968<br>(15.5) | 0.970<br>(15.8) | 1.24<br>(36.4) | 1.71<br>(47.1) | 1.20*<br>(9.58) | NC | 7.08¶<br>(154) |
| **RA<sub>AUCτ</sub>** | --- | --- | --- | --- | --- | --- | --- | 0.938<br>(7.80) | 1.13<br>(9.25) | 1.04<br>(18.0) | 0.961<br>(14.7) | 0.977<br>(11.7) | 1.16<br>(12.2) | 1.09<br>(11.8) |
| **RA<sub>Cmax</sub>** | --- | --- | --- | --- | --- | --- | --- | 0.843<br>(16.0) | 1.10<br>(11.4) | 0.969<br>(23.8) | 0.861<br>(14.3) | 0.962<br>(18.5) | 1.04<br>(20.0) | 1.09<br>(7.15) |
| **Ae<sub>0-12</sub> (mg)** | 0.391<br>(55.7) | 0.993<br>(96.9) | 1.38<br>(81.8) | 3.37<br>(57.4) | 4.57<br>(57.0) | 11.9<br>(22.9) | 22.7<br>(34.4) | 0.336<br>(64.5) | 0.915<br>(48.0) | 1.76<br>(85.5) | 3.14<br>(63.4) | 5.32<br>(47.5) | 16.4<br>(39.9) | 18.9<br>(81.6) |
| **Fe<sub>0-12</sub> (%)** | 0.993<br>(55.7) | 1.26<br>(96.9) | 0.879<br>(81.8) | 1.43<br>(57.4) | 1.45<br>(57.0) | 2.52<br>(22.9) | 3.61<br>(34.4) | 0.854<br>(64.5) | 1.16<br>(48.0) | 1.12<br>(85.5) | 1.33<br>(63.4) | 1.69<br>(47.5) | 3.48<br>(39.9) | 3.00<br>(81.6) |

\* N=5; † N=3 ¶ N=4.
Abbreviations: $Ae_{0-12}$ = amount of dose administered recovered in urine from time 0 to 12 hours postdose; $AUC_{\tau}$ = area under the plasma concentration-time curve during a dosing interval; $AUC_{\inf}$ = area under the plasma concentration-time curve from time 0 extrapolated to infinity; $AUC_{\text{last}}$ = area under the plasma concentration-time curve from time 0 to the last measurable non-zero concentration; $C_{\max}$ = maximum observed concentration; $Fe_{0-12}$ = percentage of the dose administered recovered in urine from time 0 to 12 hours postdose; MR = metabolite ratio; N = number of subjects; NC = not calculated; RA = accumulation ratio; $t_{1/2}$ = apparent terminal elimination half-life; $t_{\max}$ = time of the maximum observed concentration.

**Figure 2.**
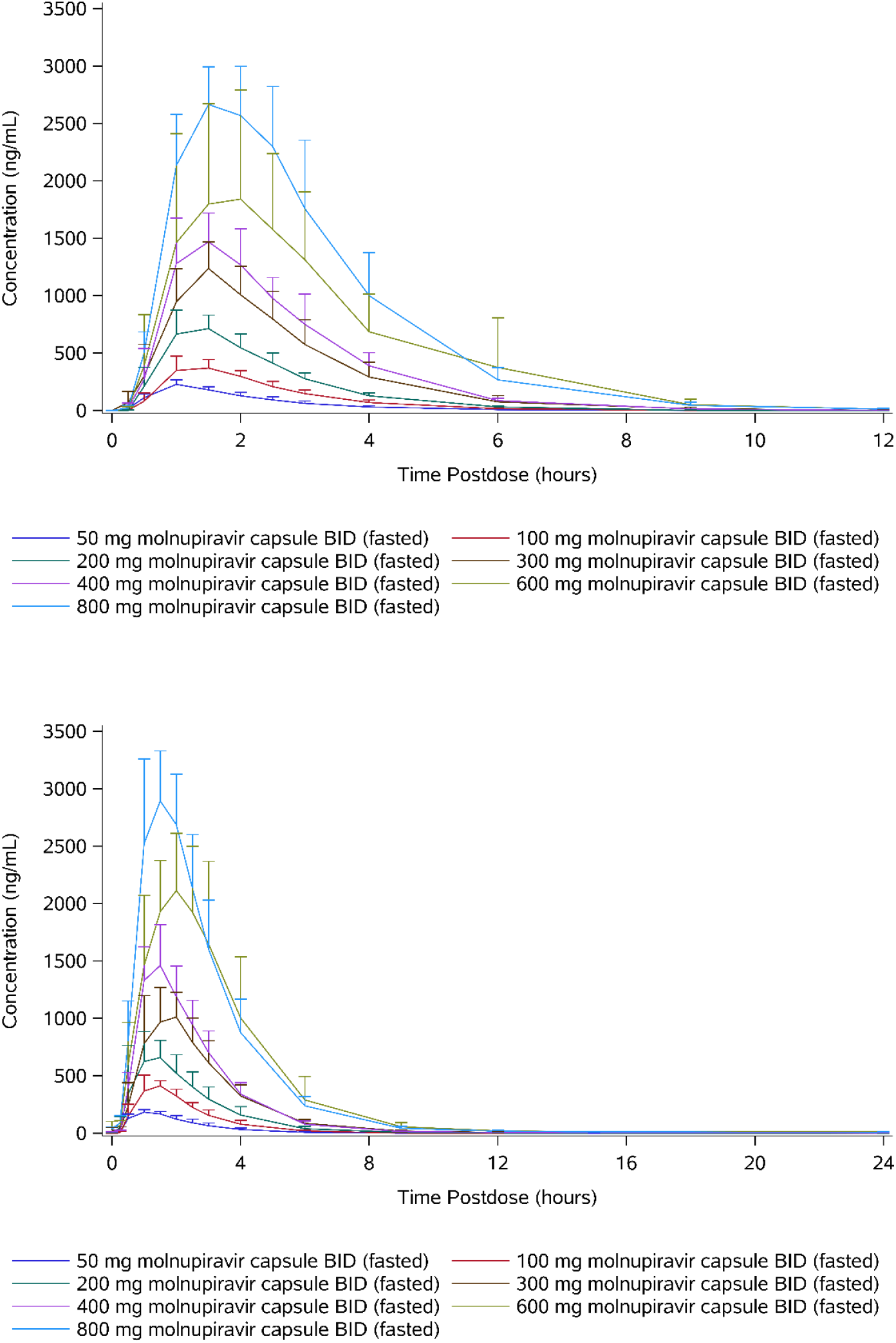
Arithmetic mean plasma concentrations of EIDD-1931 (50-800 mg molnupiravir twice-daily multiple ascending doses) on Day 1 (top) and Day 6 (bottom).

There was no evidence of accumulation, with the geometric mean accumulation ratios based on area under the plasma concentration-time curve during a dosing interval (AUC_τ_) and C_max_ between 0.938 and 1.16, and between 0.843 and 1.10, respectively, across all dose levels.

On Day 1, when assessed using the power model, mean C_max_ and AUC_inf_ increased in an approximately dose-proportional manner. However, the upper bound of the 90% confidence interval for C_max_ was slightly below unity and the lower bound of the 90% confidence interval for AUC_inf_ was slightly above unity (Table 7). On Day 6, mean C_max_ increased in a dose-proportional manner, with the 90% confidence interval containing unity. Similarly, mean AUC_τ_ increased in an approximately dose-proportional manner; however, the lower bound of the 90% confidence interval was slightly above unity (Table 7).

**Table 7.** Dose proportionality of EIDD-1931 (50-1600 mg molnupiravir single ascending doses [Day 1] and 50-800 mg molnupiravir twice-daily multiple ascending doses [Day 6])Abbreviations: AUC_τ_ = area under the plasma concentration-time curve during a dosing interval; AUC_inf_ = area under the plasma concentration-time curve from time 0 extrapolated to infinity; AUC_last_ = area under the plasma concentration-time curve from time 0 to the last measurable non-zero concentration; C_max_ = maximum observed concentration; n = number of observations.

| Parameter | Day 1 |  |  |  | Day 6 |  |  |  |
| --- | --- | --- | --- | --- | --- | --- | --- | --- |
|  | n | Slope<br>(90%<br>Confidence<br>Interval) | Between-subject<br>Geometric<br>Coefficient of<br>Variation | Lack of<br>Fit<br>P-value | n | Slope<br>(90%<br>Confidence<br>Interval) | Between-subject<br>Geometric<br>Coefficient of<br>Variation | Lack of<br>Fit<br>P-value |
| AUC<sub>inf</sub><br>(ng.h/mL) | 53 | 1.10<br>(1.06, 1.14) | 19.6 | 0.3149 | --- | --- | --- | --- |
| AUC<sub>last</sub><br>(ng.h/mL) | 54 | 1.11<br>(1.07, 1.15) | 20.8 | 0.4483 | --- | --- | --- | --- |
| AUC<sub>τ</sub><br>(ng.h/mL) | --- | --- | --- | --- | 41 | 1.08<br>(1.02, 1.14) | 20.5 | 0.3670 |
| C<sub>max</sub><br>(ng/mL) | 54 | 0.957<br>(0.916, 0.998) | 20.1 | 0.7011 | 41 | 0.971 (0.915,<br>1.03) | 20.3 | 0.7798 |

AUC_inf_ on Day 1 for the multiple-dose cohorts, where a capsule formulation was administered, was similar to those for the corresponding single-dose cohorts where a solution formulation was administered, with geometric mean ratios of between 0.91 and 1.09. Geometric mean C_max_ was slightly lower following dosing with the capsule formulation, with geometric mean ratios of between 0.76 and 1.00, and a trend to smaller ratios at higher doses. Median t_max_ occurred up to 0.75 hours later following administration of the capsule formulation, with the difference being greatest at doses ≥600 mg BID. Thus, it appears that the extent of absorption is similar for the solution and capsule formulations, but the rate of absorption appears to be slightly slower for the capsules. However, these data should be interpreted with caution because this was not a crossover study.

Between 0.854% and 3.61% of the dose was excreted in urine as EIDD-1931 on both Days 1 and 6, and, similar to single doses, the majority was excreted in the first 4 hours postdose (Table 6). There was no consistent dose-related trend in the percentage of dose administered recovered in urine during a dosing interval (Fe_0-τ_) or CL_R_ at doses ≤200 mg BID. However, there was a trend for Fe_0-τ_ and CL_R_ to increase with dose at doses >200 mg BID, with a 4-fold increase in dose from 200 to 800 mg BID resulting in a 16-fold increase in the amount of the dose recovered in urine during a dosing interval (Ae_0-τ_) on Day 1 and an 11-fold increase in Ae_0-τ_ on Day 6.

#### (iii) Food effect

Concentrations of molnupiravir were generally not quantifiable and pharmacokinetic parameters were not calculable. Concentrations of EIDD-1931 were quantifiable at 0.25 hours postdose for 2 subjects in the fasted state, but no subjects in the fed state. The first quantifiable concentrations in the fed state were between 0.5 and 1.5 hours postdose.

Following administration of 200 mg molnupiravir in the fed state, t_max_ of EIDD-1931 occurred later, with a median of 3.00 hours postdose versus 1.00 hour postdose (Table 8 and Figure 3). Generally, the slower absorption and later t_max_ in the fed state was reflected in lower C_max_; however, 1 subject had similar profiles for both treatments (data not shown). Mean C_max_ was approximately 36% lower in the fed state compared to the fasted state, but exposure (assessed by AUC_inf_) was similar for both fed and fasted states and demonstrated that the extent of absorption was similar. Following C_max_, concentrations of EIDD-1931 declined in an essentially monophasic manner in both the fed and fasted state and remained quantifiable until between 9 and 15 hours postdose in the fed state and between 6 and 9 hours in the fasted state. The mean t_1/2_ was similar between fed and fasted treatments, with values of 1.09 and 0.977 hours, respectively. Urine pharmacokinetic parameters were similar to those reported for single ascending doses.

**Table 8.** Pharmacokinetic parameters of EIDD-1931 (food effect) Geometric means (percentage coefficient of variation) are presented, with the exception of t_max_where medians (minimum – maximum) are presented.

| Parameter | 200 mg Fasted<br>(N = 10) | 200 mg Fed<br>(N = 10) |
| --- | --- | --- |
| AUC<sub>last</sub> (ng.h/mL) | 1950 (29.9) | 1870 (29.0) |
| AUC<sub>inf</sub> (ng.h/mL) | 1980 (29.3) | 1890 (28.8) |
| C<sub>max</sub> (ng/mL) | 893 (36.8) | 575 (27.7) |
| t<sub>max</sub> (h) | 1.00 (1.00 – 2.50) | 3.00 (2.00-4.00) |
| t<sub>1/2</sub> (h) | 0.977 (13.0) | 1.09 (16.3) |
| FE<sub>AUCinf</sub> | --- | 0.955 (9.7) |
| FE<sub>Cmax</sub> | --- | 0.644 (22.5) |
| Ae<sub>0-24</sub> (mg) | 1.76 (45.6) | 1.63 (54.2) |
| Fe<sub>0-24</sub> (%) | 1.12 (45.6) | 1.04 (54.2) |

Abbreviations: Ae<sub>0-24</sub> = amount of the dose administered recovered in urine from time 0 to 24 hours postdose; AUC<sub>inf</sub> = area under the plasma concentration-time curve from time 0 extrapolated to infinity; AUC<sub>last</sub> = area under the plasma concentration-time curve from time 0 to the last measurable non-zero concentration; C<sub>max</sub> = maximum observed concentration; Fe<sub>0-24</sub> = percentage of the dose administered recovered in urine from time 0 to 24 hours postdose; FE<sub>AUCinf</sub> = ratio of area under the plasma concentration-time curve from time 0 extrapolated to infinity (fed:fasted); FE<sub>Cmax</sub> = ratio of maximum observed concentration (fed:fasted); t<sub>1/2</sub> = apparent terminal elimination half-life; t<sub>max</sub> = time of the maximum observed concentration.

**Figure 3.**
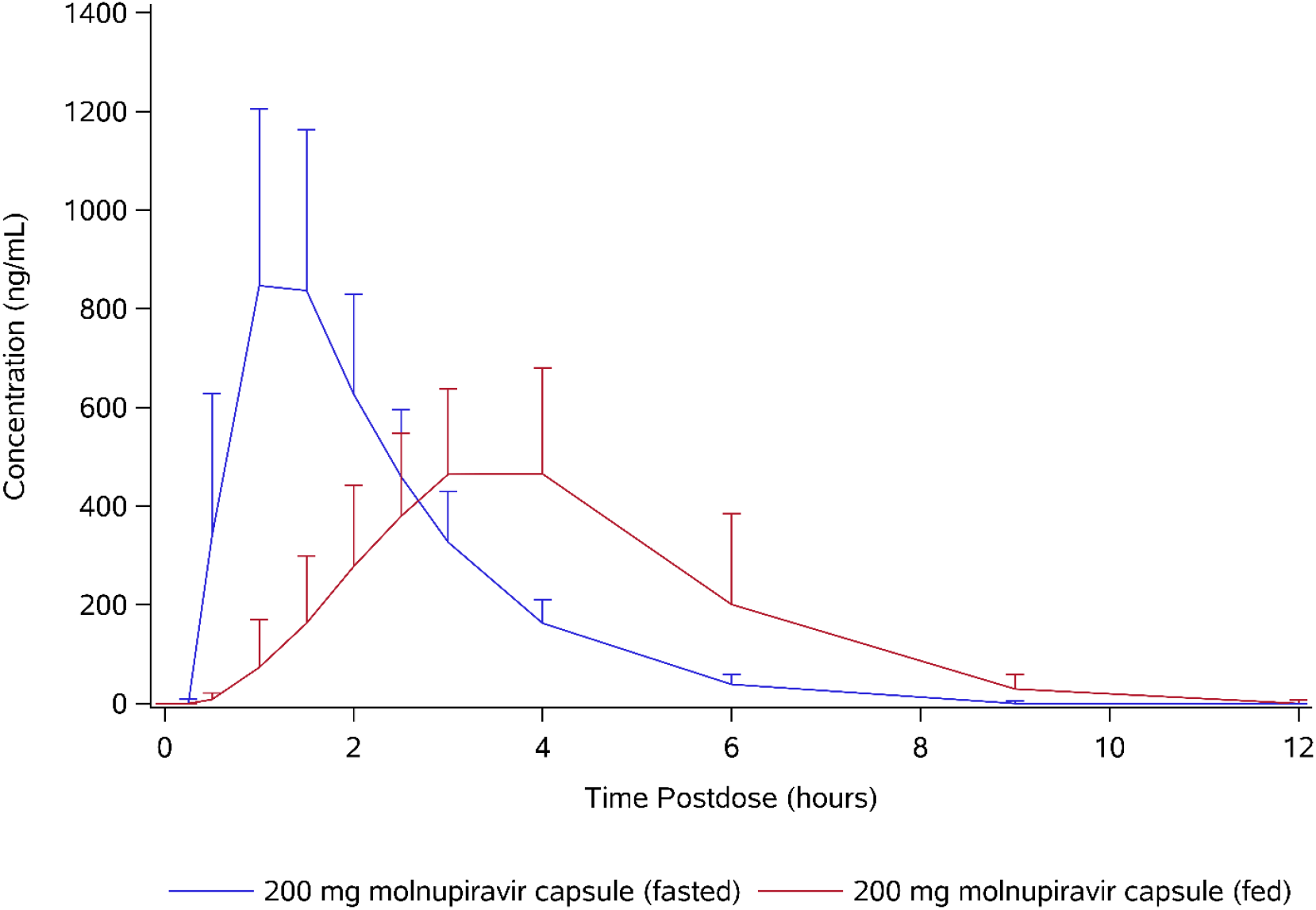
Arithmetic mean plasma concentration of EIDD-1931 (food effect).

## Discussion

There is a significant need for an antiviral drug against coronaviruses with pandemic potential that is generally safe and well tolerated and can be easily administered in the outpatient setting. The oral route of administration of molnupiravir makes it appropriate and convenient for administration to outpatients.

Additionally, if further studies demonstrate that molnupiravir has an acceptable risk/benefit profile, its oral route of administration may allow for evaluation in a prophylactic setting. Among 350 cases of COVID-19 in the United States where exposures were solicited, approximately half of respondents reported a known exposure in the 2 weeks prior to the illness, with 45% exposed from an infected family member and 34% exposed from an infected work colleague (12). This suggests a need for treatment of infected individuals and their close contacts in an outpatient setting to reduce viral shedding and infectivity, thereby reducing transmission.

Furthermore, SARS-CoV-2 RNA has been found in multiple organs at autopsy, indicating the ability to spread systemically (13). Molnupiravir is converted in plasma to EIDD-1931, which is subsequently well distributed to tissues including lung, spleen, and brain (9); therefore, it can localize to sites of replication in the treatment of systemic disease. An antiviral drug, such as molnupiravir, with broad distribution once absorbed is a desirable attribute for an emerging therapy for the treatment of COVID-19.

Plasma exposures expected to be efficacious, based on scaling from animal models of seasonal and pandemic influenza, were exceeded(11). Molnupiravir, having demonstrated good tolerability and dose-proportional pharmacokinetics with relatively low variability following administration to healthy volunteers at clinically relevant doses is well positioned to be evaluated for clinical efficacy and safety in large-scale COVID-19 studies.

## Materials and Methods

### Study population

Healthy subjects, of any ethnic origin, race, or sex, aged between 18 and 60 years, inclusive, and with a body mass index between 18 and 30 kg/m^2^, inclusive, were eligible to participate in this study. Potential subjects were screened for inclusion in the study within 28 days prior to the first dose administration after voluntarily providing informed consent.

### Study design

Eligible subjects were randomized in a 3:1 ratio to either study drug or placebo in the single- and multiple-ascending-dose parts. Placebo was chosen as the control treatment to assess whether any observed safety and tolerability effects were related to the treatment or simply reflected the study conditions. Each dose-escalation cohort comprised 8 subjects, with single oral doses of 50 to 1600 mg administered in the single-ascending-dose part and BID doses of between 50 and 800 mg for 5 days with a final dose on the morning of Day 6 being administered in the multiple-ascending-dose part. Safety and tolerability data up to 72 hours postdose (post final dose for multiple ascending doses) were reviewed prior to each dose escalation to ensure that it was safe to proceed with the planned design and multiple doses were not administered until the planned total daily dose had been shown to be safe and well tolerated as a single dose. Subjects were followed for 14 days following dose administration (post final dose for subjects who received multiple doses) for assessments of safety, tolerability, and pharmacokinetics, which exceeded 5 half-lives of EIDD-1931 observed in nonclinical studies (t_1/2_ = 9.1 hours in dogs and 5 hours in ferrets).

Subjects in the food-effect evaluation were randomized in a 1:1 ratio to either 200 mg molnupiravir in the fed state followed by 200 mg molnupiravir in the fasted state, or vice versa. Randomization was performed and subjects were followed as described for the single- and multiple-ascending-dose parts. There was a 14-day washout period between doses in the food-effect evaluation.

A capsule formulation was used in all parts of the study, with the exception of single ascending doses ≤800 mg, where an oral solution formulation was used.

The study was conducted in accordance with the International Council for Harmonisation Good Clinical Practice guidelines, the ethical principles outlined in the Declaration of Helsinki, and the European Union Clinical Trial Directive (2001/20/EC) following approval by applicable regulatory authorities and receipt of a favorable opinion by an ethics committee.

### Objectives and endpoints

The primary objectives of this study were: (i) to determine the safety and tolerability of single and multiple ascending doses of molnupiravir, and (ii) to assess the effect of food on the pharmacokinetics of molnupiravir and EIDD-1931 following a single dose. The secondary objective of the study was to define the pharmacokinetics of molnupiravir and EIDD-1931 in plasma and urine following single and multiple doses in healthy subjects. The safety and tolerability endpoints were clinical laboratory evaluations, vital signs, electrocardiograms, physical examinations, and adverse events. Pharmacokinetic endpoints included plasma and urinary parameters.

### Criteria for evaluation

Following single doses, plasma and urine samples for pharmacokinetic analysis were collected up to 72 and 48 hours postdose, respectively. Following multiple doses, plasma and urine samples were collected up to 12 hours after the first dose and up to 192 and 72 hours postdose, respectively, after the final dose. Pharmacokinetic samples were analyzed for molnupiravir and EIDD-1931 using a validated liquid chromatography with tandem mass spectrometry bioanalytical method. Pharmacokinetic parameters were calculated using noncompartmental methods in Phoenix WinNonlin Version 8.1.

Safety and tolerability were monitored throughout study participation through recording of adverse events, clinical laboratory evaluations, vital signs, electrocardiogram measurements, and physical examinations. Adverse events were monitored by observation of signs and symptoms, open questioning, and spontaneous reporting, and were graded according to the National Institutes of Health Division of Microbiology and Infectious Disease toxicity grading scale.

### Sample size and statistical analysis

No formal sample size calculation was conducted; however, 8 subjects per cohort was considered sufficient for an adequate pharmacokinetic analysis following single and multiple doses. Ten subjects in the food-effect evaluation was typical of food-effect studies with a randomized crossover design. Data were presented using descriptive statistics.

### Data availability

Certain data (for example, subject-level data) will be withheld from disclosure due to national and/or regional regulatory requirements and data protection policies. Other data will be available on request within 6 months of this publication via direct communication with the corresponding author.

## Data Availability

Not yet published.

## Acknowledgements

Ridgeback Biotherapeutics LP funded this study and has subsequently entered into a collaboration with Merck to jointly develop molnupiravir. WP is an employee of Ridgeback Biotherapeutics LP and previously was a consultant to Emory Institute of Drug Development; MM and LS are employees of Ridgeback Biotherapeutics; WH is a co-founder, owner, and advisor to Ridgeback Biotherapeutics; FA, HM, and NE are employees of Covance Clinical Research Unit Limited (the drug development division of Laboratory Corporation of America Holdings), which was responsible for the clinical conduct of this study; JB is an employee of Covance Clinical Research Unit Limited and is an equity holder of Laboratory Corporation of America Holdings; GP is the director of the Emory Institute of Drug Development, is the chief executive officer of Drug Innovation Ventures at Emory, and has a financial interest in molnupiravir.

The authors are grateful to the extraordinary collaboration between Ridgeback Biotherapeutics, Covance, the Medicines and Healthcare products Regulatory Agency, and the North East – York Research Ethics Committee that made the study conduct efficient and possible. The authors would also like to acknowledge Dr Mark Stead of Covance Medical Writing for help with the preparation of this manuscript.

